# Evaluating the Acceptability of Lyme Disease Prevention Strategies in Ottawa, Canada: A Mixed-Methods Study

**DOI:** 10.64898/2026.10.23.26366289

**Authors:** Katarina Ost, Michala Norman, Kate Zinszer, Cindy Feng, Alison Krentel, Manisha A. Kulkarni

## Abstract

**Introduction:** In recent years, the province of Ontario has reported one of the highest Lyme disease incidence rates in Canada due to the abundance of habitat that favours the survival and reproduction of ticks and wildlife species that carry the LD pathogen (*Borrelia burgdorferi*). The aim of this study was to understand the acceptability and feasibility of various methods for LD prevention in the context of peri-urban settings in Ottawa, where LD is an emerging problem.

**Methods:** This research used an exploratory sequential mixed methods design. We conducted 12 semi-structured key informant interviews with public health practitioners, land management officials, and community members to identify the most locally acceptable and feasible LD interventions. We used themes that emerged during the interviews to develop and refine a previously validated questionnaire and implement a population-representative cross-sectional survey to evaluate public acceptability and feasibility, which applied a 9-point acceptability scale to this context. The survey also collected information on frequency of tick bites, uptake of LD prevention behaviours, and explanatory variables including demographic information, property type and characteristics, and engagement with occupational or recreational outdoor activities. We distributed the survey to the Ottawa public via a web panel in 2024.

**Results:** A total of 1015 participants responded to the survey. Findings from both key informant interviews and the population survey indicated higher acceptability of personal protection strategies and landscape management strategies. Twenty-nine (29) percent of survey respondents reported knowing someone with an LD diagnosis and 3% reported having received a diagnosis of LD at some point, while 22% and 16% of respondents reported having at least one tick encounter or tick bite in the last year, respectively. Only 10% reported consistently using some methods of protection against ticks while outdoors. Acceptability of interventions was more consistently associated with respondents’ sociodemographic characteristics than with their risk and exposure indices, which varied greatly in their contribution to acceptability across intervention types.

**Conclusions:** In our study, participants generally found host-targeted and chemical strategies to be the least acceptable type of intervention to prevent LD, while personal prevention strategies, land management, and natural alternatives to chemicals were found to be the most acceptable in the Ottawa context. The factors associated with higher acceptability varied by intervention type. This information will be helpful in producing high quality, targeted messaging to promote the uptake of different LD interventions. Overall, these findings will help to better inform policy on feasible and acceptable LD prevention practices that respond to local population preferences and context.

**Ethics:** Ethical approval for this study was received from the University of Ottawa Health Sciences and Science Research Ethics Board (file number H-11-22-8400).

## 1. Introduction

Lyme disease (LD) is a tick-borne disease caused by the bacteria *Borrelia burgdorferi s.l.* and is a growing public health threat in Canada, with reported cases rising from 682 to 4,785 between 2013 and 2023 (1). In Ottawa, the national capital city, the incidence of LD has grown from 7.7 to 37 cases per 100,000 people between 2015 and 2024 (2). Previous studies in Ottawa have identified several hot spots of LD risk due to the city’s particular combination of land use, urban expansion, and climatic/ecological suitability, and presence of the primary vector tick in this region (*Ixodes scapularis*) (3–6).

Strategies to prevent LD can be broadly grouped into four categories. The first group comprises environmentally applied strategies, which include chemical, biological, and natural tick control strategies. These products are most commonly applied to the ground or vegetation as a spray or pellet formulation with the end goal of killing or repelling tick vector species in a certain area (7–12). The second group are host-targeted strategies, which may target the reproductive host of *I. scapularis* ticks, the white-tailed deer (*Odocoileus virginianus*), in an effort to reduce the overall population of ticks, or prevent the deer from moving through human use areas and transporting ticks into these spaces (8,13). These strategies may also target the disease amplification hosts, primarily the white footed mouse (*Peromyscus leucopus*), with the primary aim of reducing the prevalence of the *B. burgdorferi* bacteria infection in ticks and their hosts (14,15). The third group includes landscape modification strategies, which comprise options like ecotone modification and vegetation management to reduce tick habitat around human use areas (7). Finally, the focus of many LD prevention campaigns is on personal protection methods, such as using repellent sprays containing DEET or picaridin on the skin, permethrin-treated clothing, or wearing long clothing and tucking pants into socks etc. (16).

Knowledge, Attitudes, and Practices (KAP) surveys are commonly used in cross-sectional studies to assess what populations know about a disease, how they perceive associated risks, and which preventive behaviours they adopt (17). In Ottawa, a KAP survey was previously conducted to examine residents’ awareness and perceptions of ticks and Lyme disease, as well as the strategies they use to prevent tick encounters and tick bites, and some of their risk behaviours (18). Though KAP surveys provide valuable insight into awareness and behaviours, there is continued need to understand how a given community would respond to specific disease prevention or control interventions to inform the implementation of effective public health strategies. As a next step, this acceptability and feasibility study focused directly on the conditions required for successful and sustainable interventions, recognizing that public acceptability plays an important role in the uptake of health and social interventions. Although several studies have examined acceptability of tick control or LD prevention activities in other contexts, evidence shows that acceptability is context-dependent and changes over time (19–21). This reinforces the need to further assess and understand acceptability, and its drivers, locally, in cities like Ottawa where LD is a growing public health concern.

Our overarching study aim was to assess the acceptability and feasibility of tick control and Lyme disease prevention options in Ottawa, Canada. The qualitative phase sought to explore important themes surrounding key informants’ understanding of public and organizational acceptability with regards to the interventions, which informed the development and refinement of a survey questionnaire. The quantitative phase then assessed the comparative acceptability of tick control and LD prevention options in a population-based sample using a cross-sectional survey and identified important sociodemographic and risk factors associated with high acceptability across intervention categories. We integrated both phases using an exploratory sequential mixed methods approach to provide a comprehensive understanding of the relative acceptability of LD prevention strategies amongst Ottawa residents in the current socioecological context.

## 2. Methods

### 2.1 Ethics

Ethical approval for this study was received from the University of Ottawa Health Sciences and Science Research Ethics Board (file number H-11-22-8400). This study employed an exploratory mixed-methods research design combining key informant interview (KIIs) and a population level cross-sectional survey administered through an online web panel by the online survey and analytics company Leger 360, which has expertise in collecting representative survey data in the greater Ottawa area (22). Key informants participated in a 1:1 verbal consenting conference with the researcher prior to the beginning of the interview, while survey participants consented through an online form.

### 2.2 Study design

We used an exploratory sequential mixed methods approach to measure acceptability for several tick control and LD prevention interventions in Ottawa. Phase 1 of the study included qualitative key informant interviews (KIIs), followed by Phase 2, a quantitative approach that included the development and refinement of an existing survey, and distribution of a cross-sectional survey targeting a representative sample of adults in the city of Ottawa using a stratified sampling design.

### 2.3 Phase 1-Key informant interviews

We conducted 12 semi-structured key informant interviews to assess the feasibility and acceptability of LD prevention and control approaches in the local Ottawa context and modify the subsequent questionnaire. These KIIs included participants from a variety of fields, areas of expertise and roles who were recruited using a purposive sampling frame (23). The sampling frame included field scientists, public health professionals, healthcare professionals, individuals with lived experiences of LD (including patient representatives and caregivers), and community members with experiences living or recreating in environments where tick exposure may occur. We collected information on participants’ personal or professional acceptability of various tick or Lyme disease control options, and their perceived barriers, feasibility, and public concerns (Supplemental File-interview guide). Recruitment was ongoing to the point of thematic saturation, and for each interview we collected an audio file and verbatim transcript (23). Data were analysed in NVivo Qualitative Data Analysis Software (v 14) by two researchers, who independently coded three transcripts to ensure interrater reliability and expand the codebook. The final codebook was applied to the remaining nine transcripts (24).

We used a combination of inductive and deductive coding on our transcripts. We aimed to capture overarching (inductive) themes of drivers of acceptability and feasibility (or the lack of). We also used a deductive approach, using predefined codes that identified data specifically relevant to the four pre-defined tick control and LD prevention categories (i.e. environmentally applied, host targeted, landscape management, and personal tick protection strategies). Furthermore, results emerging from the interviews informed the refinement of an existing questionnaire previously used by Logan et al. 2024, to better capture data on the acceptability of interventions that population members had direct control over, and acceptability of interventions that survey respondents would or would not find acceptable if they were implemented by a governmental body or other kind of organization (18).

### 2.4 Phase 2.1-Questionnaire adaptation and design

We modified the questionnaire used in the study by Logan et al. to include a brief description of each category of tick control or LD prevention strategies and incorporated an infographic to improve intervention understanding and respondent comprehension regarding each of the acceptability domains. We also used the thematic analysis of KIIs to refine the acceptability domains used in the survey. The initial domains included personal protection, landscape management, host-targeted interventions, and environmentally applied strategies. Based on interview feedback, several categories were subdivided: personal protection was separated into behavioural and chemical strategies; host-targeted interventions were divided into deer-targeted and small mammal targeted approaches; and environmentally applied strategies were classified as natural alternatives to chemical acaricides, biological control options, or chemical acaricide control options, while landscape management remained unchanged. This ultimately resulted in eight acceptability domain measures.

For each of the domain measures, we used a 9-item composite acceptability score which was adapted from the Intervention Rating Profile tool and previously used to evaluate the acceptability of public health interventions across multiple cultural and global health contexts, and modified this to the local context and public health issue of LD (Table 1) (25–27). This composite scale assesses perceived effectiveness, personal approval, willingness to use and/or recommend the intervention, and perceived community benefit. Scores were calculated by summing the values of the nine indicators, each of which are graded by participants on a 4-point Likert scale without the neutral option (disagree a lot, disagree, agree, agree a lot) (25). On a summed score from a minimum of 9 to a maximum of 36, 22.5 is considered the threshold of acceptability as the median score (25).

**Table 1:** Nine indicator acceptability metric adapted from Krentel et al 2021 (25)

| *Number* | *Indicator* |
| --- | --- |
| 1 | The strategies in this category work against ticks |
| 2 | The strategies in this category work against Lyme disease |
| 3 | I would use the strategies in this category on my own property (or if I had a property) |
| 4 | I would like the strategies in this category to be used in public spaces |
| 5 | I would recommend the strategies in this category to friends or family |
| 6 | I would be willing to change the way I take care of my property (or if I had a property) to incorporate the strategies in this category |
| 7 | I like the strategies in this category |
| 8 | The strategies in this category are a good way to improve health and prevent illness caused by ticks and tick-borne diseases |
| 9 | Overall, these strategies will help my community |
| *Modified from (Krentel et al. 2021) (25)* |  |
| *Participants answered along the continuum of disagree a lot, disagree, agree, agree a lot.* |  |

The survey was piloted by five individuals, (three research assistants, and two members of the public). Two of these individuals (one public member and one researcher) validated the translation into French. The survey was then pre-tested with a sample of 15 web panel members to assess response functionality, and data capture prior to full questionnaire deployment. The cross-sectional online survey of adults residing in Ottawa, Canada was distributed on November 13^th^ - 26^th^, 2024 through Leger 360, an online survey and analytics company. Leger invited a representative sample of participants from their panel of Ottawa residents using a recruitment script developed by the research team. Respondents were able to complete the survey in either English or French, the two official languages of the region. The quantitative survey dataset provided to the research team contained unique participant identifiers but no direct personal identifiers, such as names or dates of birth.

We used a quota-based, non-probability stratified sampling strategy to ensure geographic representation across Ottawa’s major residential zones (Urban, Rural, Suburban West, Suburban South, and Suburban East (Fig. 1), similar to the sampling approach used by Logan et al. 2024 (28). These strata definitions were created from the previous study using population density, walkability, and car-commuting rates from the Ottawa Neighbourhood Study (ONS), and are described in detail by Logan et al. 2024 (28). The strata broadly correspond to different levels of LD environmental risk, with higher environmental risk in the rural and suburban strata compared to the urban zone (29). Furthermore, the original stratification used the Ottawa Greenbelt, which separates the urban core from surrounding suburban zones, as a geographic divider. In our study, we applied the same general stratification concept by assigning participants to strata based on the Forward Sortation Areas (FSAs) of their home postal code. This allowed us to map individuals to the five settlement types and maintain balanced sampling across urban, rural, and suburban neighbourhoods. We aimed to collect 200 responses per strata in order to allow us to estimate at least 1.67 odds of Ottawa respondents having high acceptability of each intervention with 80% power, 95% confidence and 5% margin of error (calculated in G*Power 3.1)(30).

**Figure 1:**
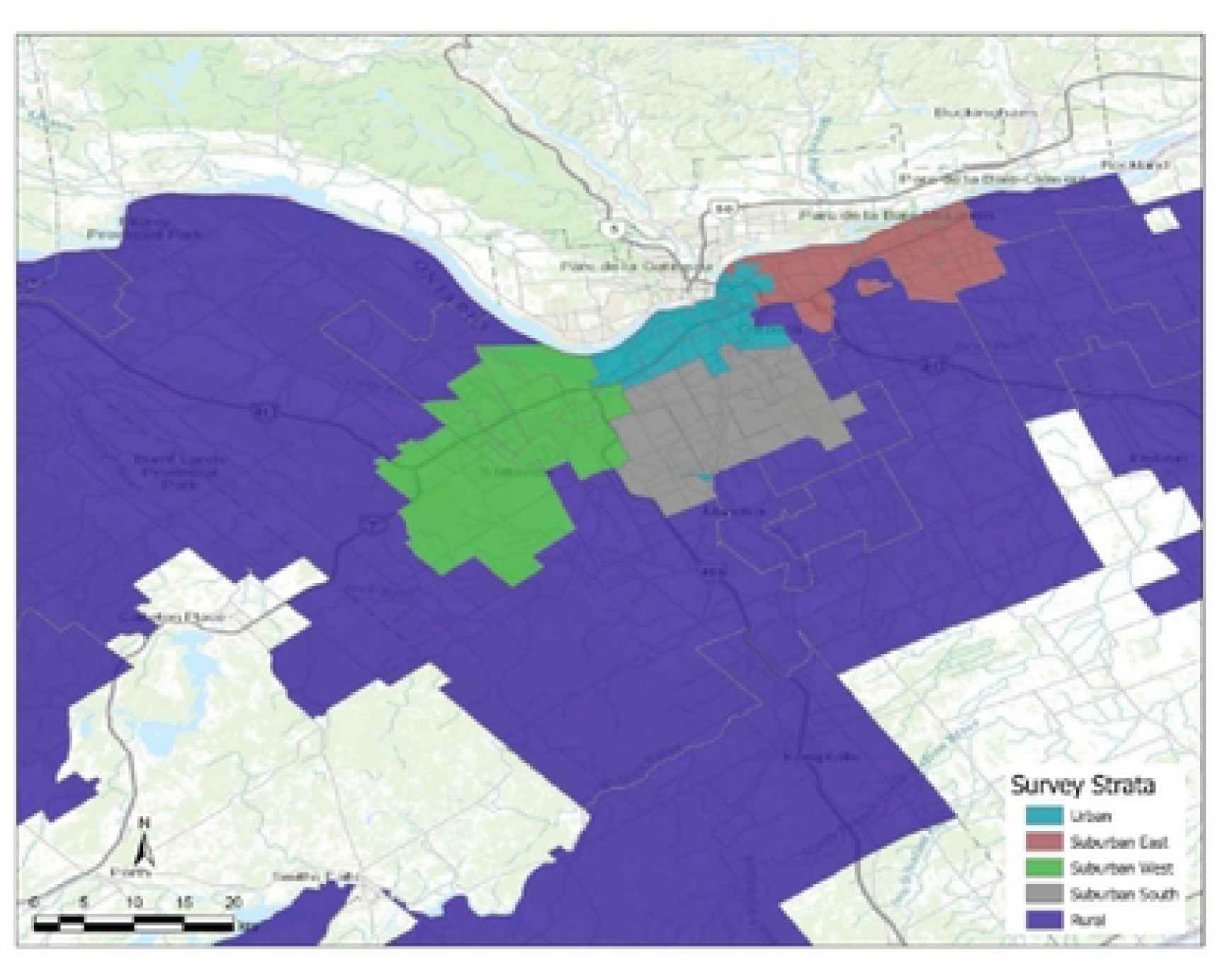
Classification of Ottawa neighborhood by postal code for survey distribution across five sampling strata: Green-Suburban West, Blue-Urban, Grey-Suburban South, Red-Suburban East and Purple-Rural (28).

The questionnaire captured data on sociodemographic variables (sex, gender, age, ethnicity, region of residence, length of residence in Canada, income, education), and risk and exposure variables (occupation - outdoor or indoor, suspected or confirmed previous LD diagnosis, yard maintenance requirements (whether participants had access to a yard and whether they were responsible for its upkeep), forest visitation frequency, pet ownership, tick encounter frequency, and tick bite frequency). The questionnaire also collected data on perceived cost, feasibility, and opinions on who should be responsible for intervention implementation.

### 2.5 Phase 2.2-Population survey statistical analysis

To evaluate predictors of intervention acceptability, we used a staged analytical aproach that combined descriptive analyses, domain-specific model development, and AIC-based model comparison. The overal analytical framework guiding this process is illustrated in Fig. 2.

**Figure 2:**
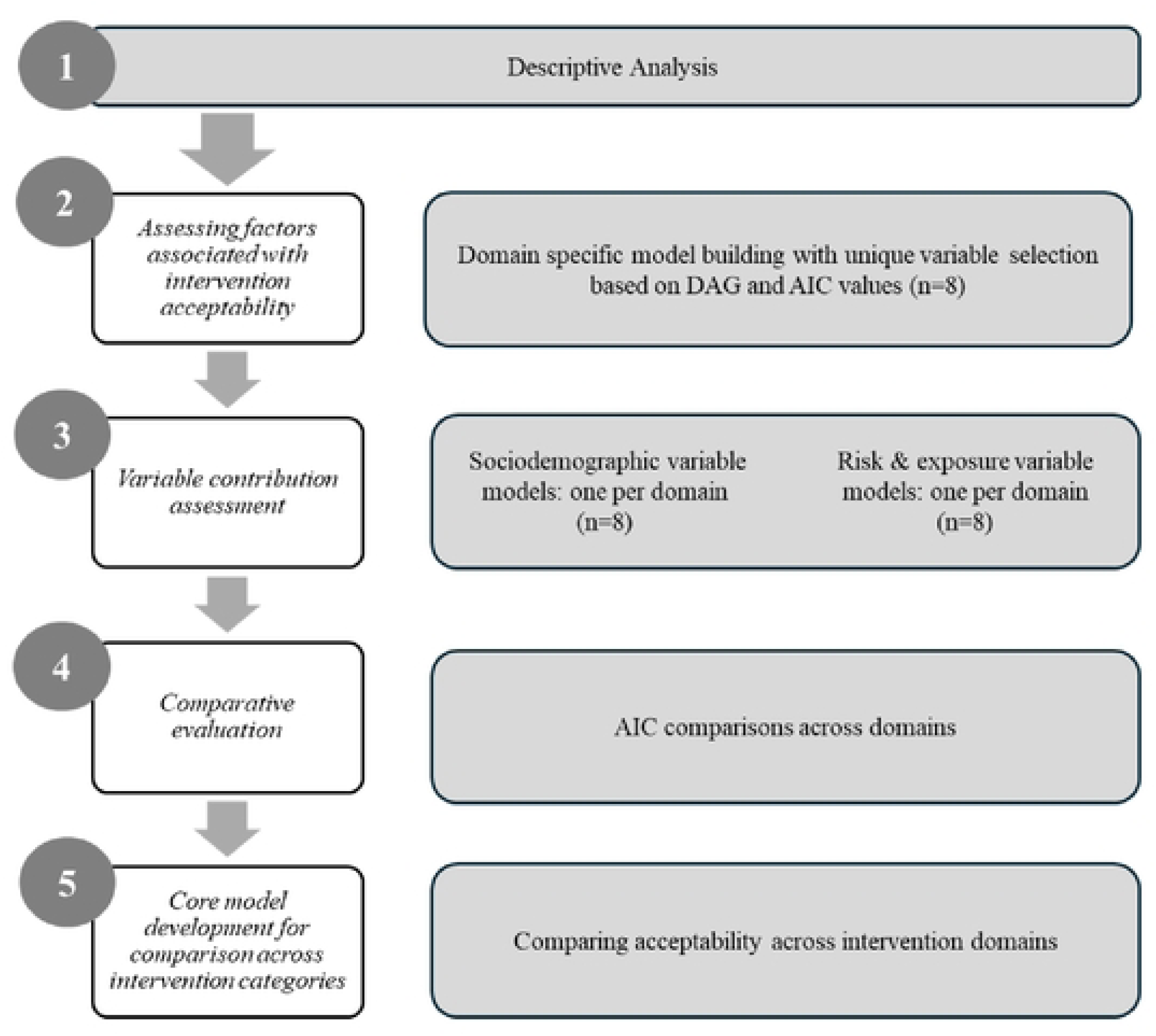
Analytical strategy for acceptability model development and analysis of intervention acceptability

#### Descriptive analysis

We first generated descriptive summaries to assess the number and proportion of respondents corresponding to the included variable categories for each section of the questionnaire. No missing data were possible for closed-ended survey items used in this analysis, as responses were required. Write in “other” responses were reviewed and reclassified into existing categories where appropriate. “Don’t know” responses were retained as a separate category or grouped with “not applicable” where conceptually appropriate. Data on ethnicity were initially collected using detailed categorical options and subsequently collapsed into a three-level variable (White, Indigenous, Other) for comparability with previous work in this region (28). Data were collected on sex and gender; however, due to collinearity between these variables, only gender was retained in the analysis. We compared survey respondent characteristics with the 2021 Ottawa Census to assess sample representativeness and identify potential sources of selection bias using the 2021 Ottawa Census as a reference. The primary analysis subsequently focused on estimating the overall acceptability scores based on the 9-item composite acceptability score described previously and identifying factors associated with high acceptability for each of the eight primary intervention categories. Because acceptability scores were expressed as binary outcomes (e.g., acceptable >=22.5 vs. low acceptability <22.5), we fit separate multivariable logistic regression models for each intervention domain.

#### Assessing factors associated with intervention acceptability

Variable inclusion for each intervention specific model was guided by a directed acyclic graph (DAG) of hypothesized relationships amongst exposure and outcome variables. The DAG informed the set of justified covariates, including key sociodemographic variables retained a priori. Akaike’s Information Criterion (AIC) was applied to compare candidate model specifications within this framework and assess variable contribution. We compared the full model with a series of reduced models using a leave-one-out AIC approach, in which variables were removed one at a time from the full model to evaluate their relative contribution to model fit based on changes in AIC (31). We developed separate models using either sociodemographic factors (e.g. gender, age, ethnicity, region of residence, length of residence in Canada, income, education level, and primary language) or risk and exposure factors (e.g. tick encounter or bite frequency, outdoor occupation, suspected or diagnosed LD, yard access and maintenance, forest visitation frequency, pet ownership) to examine their respective associations with intervention acceptability, with cluster robust standard errors calculated, accounting for region of residence for all models (i.e., Urban, Suburban East, Suburban South, Suburban West, and Rural) (32).

#### Contribution of sociodemographic and risk/exposure variables

In order to explore the relative contribution of sociodemographic or risk and exposure variables to model fit, we compared changes in AIC values across a two model set per intervention domain consisting of a sociodemographic only model, and a risk and exposure variable only model.

Within each model set, variables were dropped in a “leave one out” method to assess their relative contribution to the model’s fit. We classified predictors associated with an increase in AIC of 2 - 4 points as having a moderate contribution to model fit, and those associated with an increase of <2 points as having minimal impact on model fit (31). Lower AIC values indicate better relative model fit, while increases in AIC following variable removal indicate a greater contribution of that variable to model fit (31).

#### Core model development for comparison across intervention categories

The DAG and AIC comparisons were subsequently applied across the sixteen intervention-specific models (two models per domain) to inform the development of a “core” model. This model consisted of common a set of variables across the models that improved model fit by at least two AIC units and were supported by the DAG structure (including sociodemographic variables (gender, age group, ethnicity, region of residence, length of residence in Canada, income and education level) and risk or exposure variables (outdoor occupation, tick encounter frequency in the last year, and forest visitation frequency)). We applied these core variables to a logistic regression estimated using a generalized estimating equation (GEE) to allow direct comparison of the models across intervention categories. Respondent ID was specified as the clustering variable, with an exchangeable working correlation structure to account for within-respondent correlation. All analyses were performed in Stata (version 18.5).

#### Sensitivity analysis

The acceptability cutoff score of 22.5 has been used in several previous studies as threshold for acceptability, as it is midpoint of the score range from 9-36, low to positive acceptability (25,26). Recognizing the novel application of this acceptability tool to LD and the need to examine the suitability of this cutoff score, we performed a sensitivity analysis focused on more strict thresholds (≥23.5 and ≥25) to assess whether relative patterns of acceptability were robust to these definitions (Supplemental Table 1). Lower thresholds were not explored further, as values below this midpoint would classify low acceptability as “high”, limiting conceptual interpretability. We found that most variables kept their relative acceptability classification, with the exception of environmentally applied chemicals, which had a more bimodal distribution of acceptability scores compared to the other acceptability domains; we therefore opted to apply the 22.5 cutoff threshold in our analysis. In addition, the score of 22.5 and the relative acceptability of the domains was supported by our qualitative results, indicating that this cutoff was appropriate for this analysis.

## 3. Results

### 3.1 Qualitative phase

#### 3.1.1 Characteristics of Key informants

We included twelve key informants in our study and concluded recruitment once no additional themes were generated through interviews, indicating thematic saturation (23). Key informants were from a range of backgrounds and included program managers in environmental and public health programs, as well as community members and representatives from recreational organizations (Table 2). There was a relatively even distribution of men (5) and women (7) represented amongst the key informants, and ages ranged from 20s-70s. All interviews were conducted in English.

**Table 2:** Key informant participant characteristics, including sector, role type, and relevant expertise.

| ID | Sector | Role Type | Relevant Expertise |
| --- | --- | --- | --- |
| KI1 | Natural sciences and environmental policy | Program manager | Wildlife systems, staff exposure concerns, communications |
| KI2 | Natural resource management | Scientist | Staff exposure concerns, ecological concerns and communications |
| KI3 | Recreational organization | Executive board member | Member exposure concerns |
| KI4 | Health | Program manager | Communications |
| KI5 | Environmental conservation organizations | Founder and volunteer | Communication, community exposure concerns |
| KI6 | Community association | Volunteer | Environmental work, community exposure concerns |
| KI7 | Community association | Board member | Environmental communications, community exposure concerns |
| KI8 | Recreational organization | Founder and volunteer | Member exposure concerns |
| KI9 | Health research | Researcher and LD patient | Patient community communications |
| KI10 | Health professional, member of public | Pediatric health professional and member of public | Family exposure concerns |
| KI11 | Health research | Social science researcher | Family exposure concerns |
| KI12 | Wildlife research | Scientist, LD patient | Research team exposure concerns, wildlife host behaviour and wellbeing |

#### 3.1.2 Qualitative thematic findings

We identified six themes (two inductive and four deductive) that emerged from the key informant interview coding process: unintended consequences, personal or external control, environmentally applied strategy acceptability, host targeted control acceptability, landscape management strategy acceptability, and personal protection strategy acceptability. These themes, subthemes, and example quotes are shown in Table 3.

**Table 3:** Themes and subthemes from key informant interviews.

| **Themes and subthemes**<br>**(inductive or deductive)** | **Example quote** |
| --- | --- |
| **Unintended consequences**<br>**(inductive)**<br>Concerns regarding environmental impact<br>Concerns regarding non-target species (e.g., pollinators)<br>Concerns regarding risks to pets and children | “I actually do mow to keep, some of my grass down. I probably mow more than I would otherwise to try to make sure I'm not brushing against tick stuff like leaf litter, so, from a rural landowner, maybe around the house if you're told from an ecological perspective and everything else to leave it for the bees and the things. But if you wanted to remove it away from your house... you could do that.” [Environmental conservation organization] |
| **Personal or external control and scale**<br>**(inductive)**<br>Acceptability framed by personal vs. external control<br>Feasibility influenced by implementation context<br>Scale dependent perceptions of intervention feasibility | “Can I give you answers from two different perspectives?... One group that we do interact with quite a bit and [community organization name] are the landowners up in the [peri-urban town]... We're talking rural, often multiple acres. They [rural residences] have a house. So, I think that's one group. And the other group would be the peri- rural [residences]. ...they're backing onto a natural area...so I can see there being kind of two different answers depending. Is that OK to kind of give you two different answers or would you prefer I answer for one group?” [Environmental conservation organization]<br>“For landscape management as a solution for a community... so for public spaces or for my space at home... so generally the biggest issue, and it's a little bit like when you look at climate changes generally it's you have all these stakeholders and you want them all to do the same thing. Just managing all of that [stakeholder coordination] is such a huge level of work, and because they all have their own policies and own reasons for doing stuff, and then they may not all agree. So just I think that's overarching comment for all of the solutions.” [Community association]. |
| **Environmentally applied strategy acceptability** | “I don't like the chemical control. It just doesn't sound great for the environment and like for us breathing it in, but like I also |
| **(deductive)**<br>• Low acceptability of chemical control options<br>• Greater acceptability of natural or biological alternatives<br>• Perceived high feasibility of environmental application compared to other approaches<br>• Concerns regarding frequency of reapplication and maintenance | probably don't know that much about it either. The natural alternative sounds nice.” [member of public-medical professional]<br>“I like more the more the more natural ways, the plant-based pest control and the natural enemies because as for the chemical control... we have been told in university that... their effect is usually bigger than what we expect and so yeah, I'm more comfortable with natural solutions than chemicals.” [Natural sciences and environmental policy].<br>“I mean probably they're all similarly feasible. I don't, except for obviously the natural enemies being the least feasible, but probably in terms of applying something to the environment, whether you apply [product]- like if they have to be reapplied or so I don't know enough about that.”[Health researcher] |
| **Host targeted control acceptability (deductive)**<br>• Anticipated social resistance to deer targeted strategies<br>• Interest in small mammal-targeted approaches<br>• Uncertainty regarding function and feasibility of small mammal interventions | “Deer management is a hot button topic and, we, it was discussed probably 15-20 years ago, more in terms of impacts on vegetation, and there is no appetite to talk about deer exclusion, deer reduction... And certainly, deer reduction/controlled hunts, that would be a huge public controversy.” [Natural sciences and environmental policy].<br>“I don't see that [bait boxes] as being a big a big issue. But you know people will be concerned. They would have questions- Are kids going to find these, pick them up, play with them? Those are usually the issues that come.” [Wildlife scientist] |
| **Landscape management strategy acceptability (deductive)**<br>• High acceptability of most landscape management approaches<br>• Perceived compatibility with existing recreational space management priorities<br>• Concerns regarding scalability and feasibility for individual implementation | “On wood chips [substrates], I'm going to assume they're not treated and they're just generally wood chips. I see people being super on board... What we've really discussed in the past, I think is more the vegetation clearing along the edges. So, trimming back trails so that people are not brushing up against vegetation when they're going along those trails, that's, I think the most feasible thing.... And then, we have in some cases talked about barrier approaches. So, putting down a woodchip trail instead of just a natural trail.” [Natural sciences and environmental policy].<br>“I mean, I'm thinking about it in the terms of like our farm, and what's possible there, like we do try and mow the fields where we walk through, not all but some of the fields when we go, we have a pond but try and keep that mowed for that very reason. That's probably feasible. And it's certainly, I like it. It's acceptable to me when it's done.” [Health researcher]. |
| • Low acceptability of vegetation burning |  |
| **Personal tick or LD protection strategy acceptability (deductive)**<br>• High overall acceptability of personal tick or LD protection strategies<br>• Variation in the preference across specific tick or LD prevention behaviours | “Probably they’d [our members] be more likely to use you know OFF or Deep Woods, like something with DEET because they're familiar with it. They might have a hesitancy towards permethrin... like if we can get our hands on it and it's readily available and not too expensive, providing that at the trail day, so people can apply it if they want.” [Recreational organization board member]<br>“I don't like spraying Permethrin on my clothing that I wear every day or I will dedicate a pair of pants for it. So that's what I recommend to my students as well. I do not like applying DEET or other products to my skin, directly to my skin... I would rather use protective clothing. That barrier for mosquitoes and ticks and other things than use DEET directly on my skin. I will spray it sometimes on my clothes.” [Natural sciences and environmental policy]. |

Across intervention types, participants expressed concern about unintended consequences, such as the impact of interventions on the environment, non-target species (like pollinators), pets, and children when it came to many of the interventions. Acceptability was frequently related to perceived ecological safety.

A second cross-cutting theme concerned control and responsibility over a given intervention strategy, in addition to scale and context. Participants evaluated both feasibility and acceptability based on whether an intervention fell within their direct control vs. requiring external coordination and implementation (municipal, provincial etc.). Perceived feasibility was closely tied to implementation scale and the context of responsibility.

Within deductively coded intervention categories, chemical control strategies were generally viewed unfavourably and associated with low acceptability, whereas natural or biological alternatives were viewed as being more acceptable, though participants still brought up concerns regarding environmental safety. Environmentally applied strategies were often perceived as feasible, but there were some questions regarding reapplication frequency and maintenance over time.

Similarly host-targeted interventions, specifically deer targeted strategies, faced challenges in terms of both perceived feasibility and acceptability. Professionals working in wildlife/environmental sciences and public policy consistently anticipated substantial public resistance. In contrast, small mammal strategies were interesting to participants, but participants were unclear on how they function and unsure of the feasibility.

Landscape management strategies were broadly perceived as acceptable and feasible, with the exception of vegetation burning. Ecotone modification was viewed as being well aligned with recreational space management priorities, though there were some reservations about the potential feasibility of scaling these projects to large spaces, and implementing them in a residential space.

Finally, personal protection strategies were consistently rated as acceptable; however, preferences varied across specific protective behaviours, reflecting differences in perceived personal burden, practicality, or personal health concerns.

### 3.2 Quantitative phase

#### 3.2.1 Descriptive statistics

A total of 1015 Ottawa residents participated in the survey. Participants were broadly balanced by gender, with 52.4% identifying as women and 46.7% as men, and a small proportion identifying as non-binary or other genders. Using the 2021 Ottawa Census as a reference, we found survey respondents to be generally representative of the census population in terms of geographic region, with slight overrepresentation of the urban versus rural population (Table 4). Sociodemographic characteristics were broadly balanced across categories. Most respondents fell within the 25-34 age group (18.2%), and the fewest in the 75 and older group (8.47%).

**Table 4:**
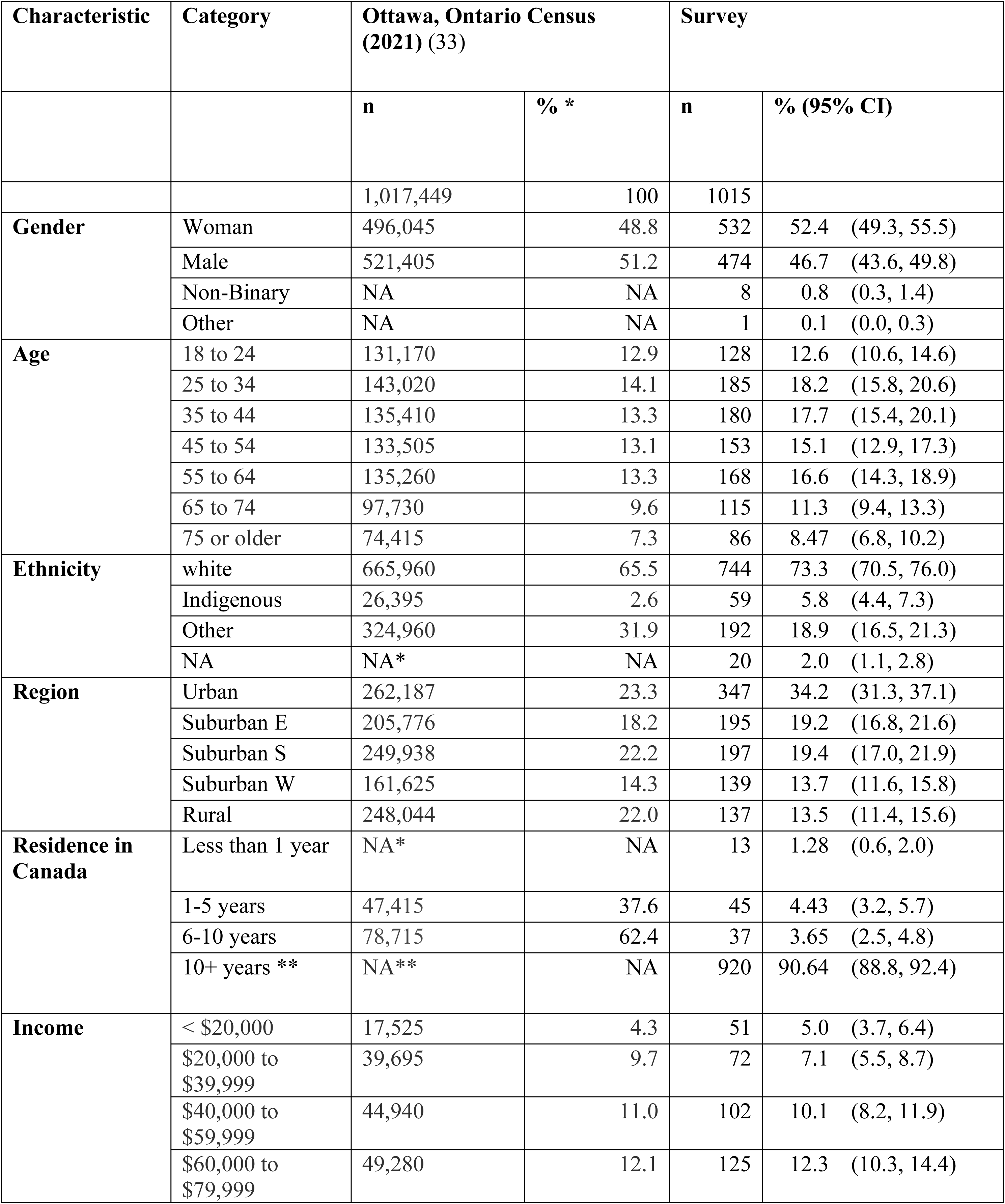
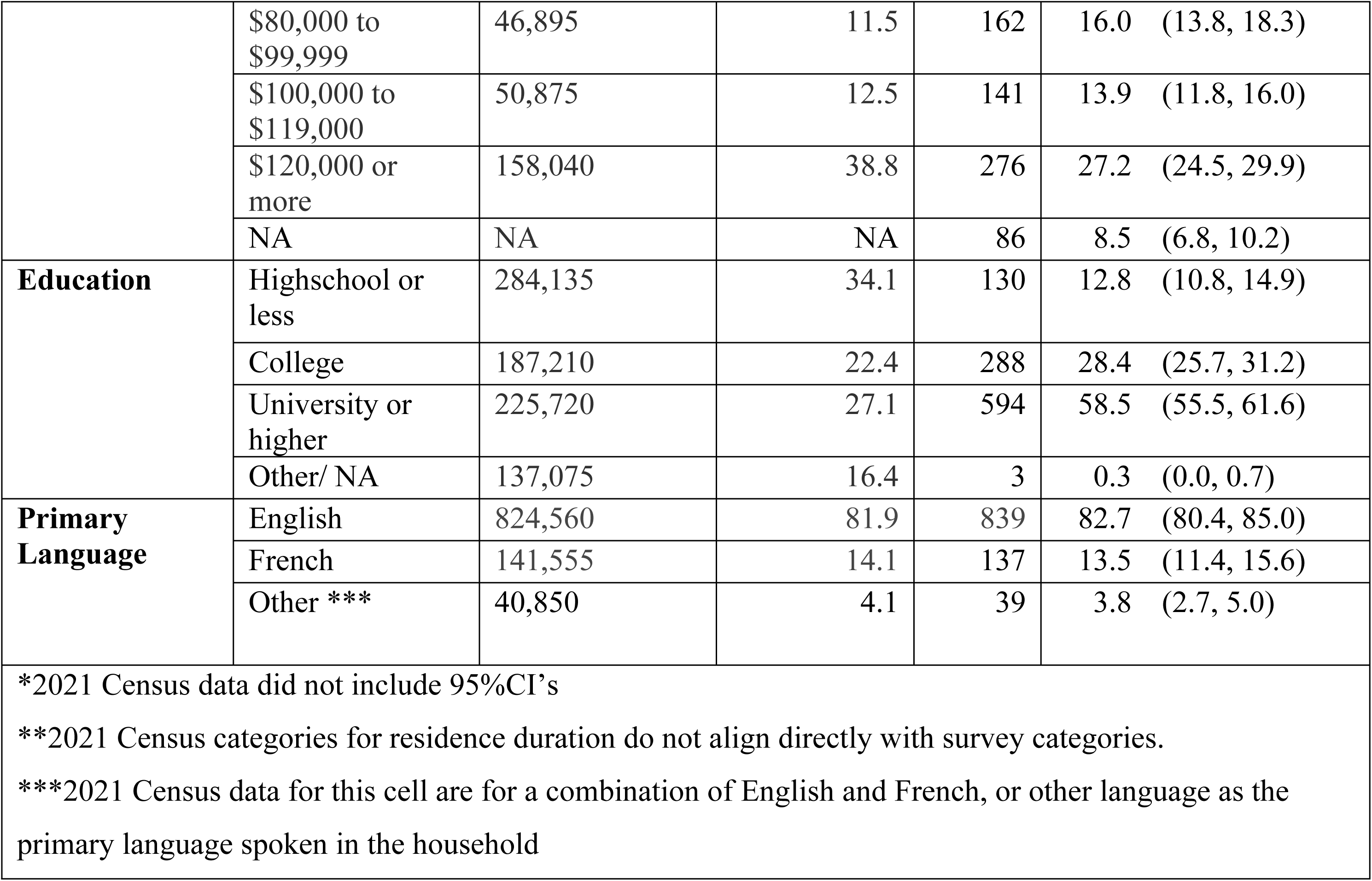
Sociodemographic characteristics of survey participants compared with the 2021 Ottawa Census.

| Characteristic | Category | Ottawa, Ontario Census<br>(2021) (33) |  | Survey |  |
| --- | --- | --- | --- | --- | --- |
|  |  | n | % * | n | % (95% CI) |
|  |  | 1,017,449 | 100 | 1015 |  |
| **Gender** | Woman | 496,045 | 48.8 | 532 | 52.4 (49.3, 55.5) |
|  | Male | 521,405 | 51.2 | 474 | 46.7 (43.6, 49.8) |
|  | Non-Binary | NA | NA | 8 | 0.8 (0.3, 1.4) |
|  | Other | NA | NA | 1 | 0.1 (0.0, 0.3) |
| **Age** | 18 to 24 | 131,170 | 12.9 | 128 | 12.6 (10.6, 14.6) |
|  | 25 to 34 | 143,020 | 14.1 | 185 | 18.2 (15.8, 20.6) |
|  | 35 to 44 | 135,410 | 13.3 | 180 | 17.7 (15.4, 20.1) |
|  | 45 to 54 | 133,505 | 13.1 | 153 | 15.1 (12.9, 17.3) |
|  | 55 to 64 | 135,260 | 13.3 | 168 | 16.6 (14.3, 18.9) |
|  | 65 to 74 | 97,730 | 9.6 | 115 | 11.3 (9.4, 13.3) |
|  | 75 or older | 74,415 | 7.3 | 86 | 8.47 (6.8, 10.2) |
| **Ethnicity** | white | 665,960 | 65.5 | 744 | 73.3 (70.5, 76.0) |
|  | Indigenous | 26,395 | 2.6 | 59 | 5.8 (4.4, 7.3) |
|  | Other | 324,960 | 31.9 | 192 | 18.9 (16.5, 21.3) |
|  | NA | NA* | NA | 20 | 2.0 (1.1, 2.8) |
| **Region** | Urban | 262,187 | 23.3 | 347 | 34.2 (31.3, 37.1) |
|  | Suburban E | 205,776 | 18.2 | 195 | 19.2 (16.8, 21.6) |
|  | Suburban S | 249,938 | 22.2 | 197 | 19.4 (17.0, 21.9) |
|  | Suburban W | 161,625 | 14.3 | 139 | 13.7 (11.6, 15.8) |
|  | Rural | 248,044 | 22.0 | 137 | 13.5 (11.4, 15.6) |
| **Residence in**<br>**Canada** | Less than 1 year | NA* | NA | 13 | 1.28 (0.6, 2.0) |
|  | 1-5 years | 47,415 | 37.6 | 45 | 4.43 (3.2, 5.7) |
|  | 6-10 years | 78,715 | 62.4 | 37 | 3.65 (2.5, 4.8) |
|  | 10+ years ** | NA** | NA | 920 | 90.64 (88.8, 92.4) |
| **Income** | < \$20,000 | 17,525 | 4.3 | 51 | 5.0 (3.7, 6.4) |
| | \$20,000 to<br>\$39,999 | 39,695 | 9.7 | 72 | 7.1 (5.5, 8.7) |
| | \$40,000 to<br>\$59,999 | 44,940 | 11.0 | 102 | 10.1 (8.2, 11.9) |
| | \$60,000 to<br>\$79,999 | 49,280 | 12.1 | 125 | 12.3 (10.3, 14.4) |
| | \$80,000 to \$99,999 | 46,895 | 11.5 | 162 | 16.0 (13.8, 18.3) |
| | \$100,000 to \$119,000 | 50,875 | 12.5 | 141 | 13.9 (11.8, 16.0) |
| | \$120,000 or more | 158,040 | 38.8 | 276 | 27.2 (24.5, 29.9) |
|  | NA | NA | NA | 86 | 8.5 (6.8, 10.2) |
| **Education** | Highschool or less | 284,135 | 34.1 | 130 | 12.8 (10.8, 14.9) |
|  | College | 187,210 | 22.4 | 288 | 28.4 (25.7, 31.2) |
|  | University or higher | 225,720 | 27.1 | 594 | 58.5 (55.5, 61.6) |
|  | Other/ NA | 137,075 | 16.4 | 3 | 0.3 (0.0, 0.7) |
| **Primary Language** | English | 824,560 | 81.9 | 839 | 82.7 (80.4, 85.0) |
|  | French | 141,555 | 14.1 | 137 | 13.5 (11.4, 15.6) |
|  | Other *** | 40,850 | 4.1 | 39 | 3.8 (2.7, 5.0) |
| *2021 Census data did not include 95%CI's<br>**2021 Census categories for residence duration do not align directly with survey categories.<br>***2021 Census data for this cell are for a combination of English and French, or other language as the primary language spoken in the household |  |  |  |  |  |

Respondents were also more likely to identify as white (73.3%), speak English as primary language (82.7%), report annual household incomes between $80,000 and $99,999 (16%), and be university educated or more (58.5%).

Overall, 29% of respondents reported knowing someone with an LD diagnosis and 3% reported having received a diagnosis of LD at some point, 22% and 16% of respondents reported having at least one tick encounter or tick bite in the last year respectively, and only 10% reported consistently using some sort of protection against ticks while outdoors (Table 5).

**Table 5:** Risk and exposure factors of participants (N=1015)

| **Risk or exposure variable** | **Category** | **N Respondents** | **% (95%CI) Respondents** |
| --- | --- | --- | --- |
| Tick encounter frequency | Never | 782 | 77.0 (74.4, 79.5) |
|  | 1-2 times per year | 183 | 18.0 (15.8, 20.5) |
|  | 3+ times per year | 50 | 4.9 (3.8, 6.4) |
| Tick bite frequency | Never | 848 | 83.5 (81.1, 85.7) |
|  | 1-2 times per year | 148 | 14.6 (12.5, 16.9) |
|  | 3+ times per year | 19 | 1.9 (1.2, 2.9) |
| Ever thought they had LD | Yes | 48 | 4.7 (3.6, 6.2) |
|  | No | 967 | 95.3 (93.8, 96.4) |
| Ever diagnosed with LD | Yes | 33 | 3.3 (2.4, 4.7) |
|  | No | 967 | 95.3 (93.8, 96.4) |
|  | I don't know | 14 | 1.4 (0.8, 2.3) |
| Know someone with LD | Yes | 293 | 28.9 (26.2, 31.7) |
|  | No | 678 | 66.8 (63.8, 69.6) |
|  | I don't know | 44 | 4.3 (3.2, 5.8) |
| Frequency of personal tick protection use | Never | 313 | 30.8 (28.1, 33.8) |
|  | Sometimes | 358 | 35.3 (32.4, 38.3) |
|  | Often | 228 | 22.5 (20.0, 25.1) |
|  | Always | 97 | 9.6 (7.9, 11.5) |
|  | I don't know | 19 | 1.9 (1.2, 2.9) |
| Yard access and maintenance | No access | 171 | 16.8 (14.7, 19.3) |
|  | Access but no maintenance | 250 | 24.6 (22.1, 27.4) |
|  | Access and maintenance | 594 | 58.5 (55.5, 61.5) |
| Yard tick prevention use | Yes, yard prevention use | 220 | 21.7 (19.2, 24.3) |
|  | No yard prevention use | 618 | 60.9 (57.8, 63.8) |
|  | No yard/No maintenance | 120 | 11.8 (10.0, 14.0) |
|  | I don't know | 57 | 5.6 (4.4, 7.2) |
| Forest use frequency | 25+times per year | 145 | 14.3 (12.3, 16.6) |
|  | 11-24 times per year | 226 | 22.3 (19.8, 24.9) |
|  | 3-10 times per year | 302 | 29.8 (27.0, 32.6) |
|  | 2 or less times per year | 231 | 22.8 (20.3, 25.4) |
|  | Never | 111 | 10.9 (9.2, 13.0) |
| Pet ownership | Yes | 393 | 38.7 (35.8, 41.8) |
|  | No | 622 | 61.3 (58.2, 64.2) |
| Pet tick or LD protection frequency | Never | 91 | 9.0 (7.4, 10.9) |
|  | Sometimes | 95 | 9.4 (7.7, 11.3) |
|  | Often | 138 | 13.6 (11.6, 15.8) |
|  | Always | 195 | 19.2 (16.9, 21.8) |
|  | No pet/ Don't know | 496 | 48.9 (45.8, 52.0) |

#### 3.2.2 Acceptability domain-specific models

Across all environmentally applied intervention types (chemical, natural, and biological), older age groups generally had lower odds of high acceptability compared with respondents aged 18-24 years (ORs: 0.33-0.67, p-values<0.02), with the exception of respondents over 75 years for biological interventions. Region of residence was also associated with acceptability across all three intervention types, although the direction of association varied by strategy. For chemical control, residents of Suburban East and rural areas had higher odds of acceptability compared with urban residents (OR: 1.77; 95% CI: 1.64–1.92; p<0.0001 and OR: 1.26; 95% CI: 1.09–1.45; p<0.001, respectively). In contrast, residents of Suburban South, Suburban West, and rural areas had lower odds of high acceptability for natural products (OR: 0.84: 95% CI 0.77, 0.91; p-value <0.001, OR: 0.71; 95% CI 0.66, 0.77; p-value <0.001, OR: 0.76; 95%CI 0.62, 0.93 p-value <0.05 respectively), while residents of Suburban South and rural areas had lower odds of high acceptability for biological interventions (95% CI: 0.64, 0.87; p-value <0.001; and 95% CI: 0.54,0.96; p-value<0.05 respectively) (Table 7).

**Table 7:** Multivariable Logistic regression models for predictors significantly associated with high acceptability of Lyme Disease prevention or tick control interventions, with number of respondents in each category, and proportion of respondents finding the intervention to be acceptable (full models in Supplemental Table 2). Variable categories showing statistically significant associations (p-value<0.05) are highlighted in green.

| **Environmentally Applied: Chemical Acceptability** |  |  |  |  |  |
| --- | --- | --- | --- | --- | --- |
|  | **Category** | **No. respondents in category** | **Proportion of respondents with high acceptability [95%CI]** | **OR [95%CI]** | **p-value** |
| **Gender** | Woman | 532 | 47.7 [0.43, 0.52] | Reference | . |
|  | Male | 474 | 67.1 [0.63, 0.71] | 2.36 [1.97, 2.82] | **<0.001** |
|  | Non-Binary | 8 | 50.0 [0.16, 0.84] | 1.12 [0.47, 2.69] | 0.8 |
|  | Other | 1 | 0.0 [0.00, 0.98] | . | . |
| **Age** | 18 to 24 | 128 | 75.0 [0.67, 0.82] | Reference | . |
|  | 25 to 34 | 185 | 55.7 [0.48, 0.63] | 0.52 [0.37, 0.73] | **<0.001** |
|  | 35 to 44 | 180 | 57.2 [0.50, 0.65] | 0.45 [0.33, 0.62] | **<0.001** |
|  | 45 to 54 | 153 | 53.6 [0.45, 0.62] | 0.42 [0.25, 0.72] | **<0.001** |
|  | 55 to 64 | 168 | 47.6 [0.40, 0.55] | 0.34 [0.22, 0.54] | **<0.001** |
|  | 65 to 74 | 115 | 53.9 [0.44, 0.63] | 0.55 [0.33, 0.92] | **0.02** |
|  | 75 or older | 86 | 58.1 [0.47, 0.69] | 0.67 [0.51, 0.89] | **0.01** |
| **Region** | Urban | 347 | 56.2 [0.51, 0.61] | Reference | . |
|  | Suburban E | 195 | 67.2 [0.60, 0.74] | 1.77 [1.64, 1.92] | **<0.001** |
|  | Suburban S | 197 | 50.3 [0.43, 0.57] | 0.79 [0.69, 0.91] | **<0.001** |
|  | Suburban W | 139 | 53.2 [0.45, 0.62] | 1.11 [0.95, 1.3] | 0.18 |
|  | Rural | 137 | 56.2 [0.47, 0.65] | 1.26 [1.09, 1.45] | **<0.001** |
| **Income** | < \$20,000 | 51 | 45.1 [0.31, 0.60] | Reference | . |
| | \$20,000 to \$39,999 | 72 | 56.9 [0.45, 0.69] | 2.03 [0.69, 5.99] | 0.2 |
| | \$40,000 to \$59,999 | 102 | 58.8 [0.49, 0.68] | 2.21 [1.18, 4.15] | **0.01** |
| | \$60,000 to \$79,999 | 125 | 54.4 [0.45, 0.63] | 1.30 [0.57, 2.94] | 0.53 |
| | \$80,000 to \$99,999 | 162 | 66.0 [0.58, 0.73] | 1.77 [0.92, 3.43] | 0.09 |
| | \$100,000 to \$119,000 | 141 | 57.4 [0.49, 0.66] | 1.45 [0.71, 2.97] | 0.31 |
| | \$120,000 or more | 276 | 56.2 [0.50, 0.62] | 1.82 [1.03, 3.22] | 0.04 |
|  | NA | 86 | 47.7 [0.37, 0.59] | 1.70 [0.9, 3.24] | 0.1 |
| Tick bite | Never | 848 | 51.3 [0.48, 0.55] | Reference | . |
|  | 1-2 times | 148 | 83.8 [0.77, 0.89] | 3.54 [2.19, 5.71] | <0.001 |
|  | 3+ times | 19 | 89.5 [0.67, 0.99] | 4.73 [1.25, 17.81] | 0.02 |
| **Environmentally Applied: Natural Acceptability** |  |  |  |  |  |
|  | **Category** | **No. respondents in category** | **Proportion of respondents with high acceptability [95%CI]** | **OR [95%CI]** | **p-value** |
| Gender | Woman | 532 | 85.9 [0.83, 0.89] | Reference | . |
|  | Male | 474 | 85.0 [0.81, 0.88] | 1.08 [0.78, 1.49] | 0.65 |
|  | Non-Binary | 8 | 75.0 [0.35, 0.97] | 0.20 [0.1, 0.41] | <0.001 |
|  | Other | 1 | 100.0 [0.03, 1.00] | . | . |
| Age | 18 to 24 | 128 | 93.8 [0.88, 0.97] | Reference | . |
|  | 25 to 34 | 185 | 88.1 [0.83, 0.92] | 0.48 [0.38, 0.62] | <0.001 |
|  | 35 to 44 | 180 | 84.4 [0.78, 0.89] | 0.36 [0.28, 0.46] | <0.001 |
|  | 45 to 54 | 153 | 85.0 [0.78, 0.90] | 0.41 [0.26, 0.64] | <0.001 |
|  | 55 to 64 | 168 | 80.4 [0.74, 0.86] | 0.33 [0.19, 0.55] | <0.001 |
|  | 65 to 74 | 115 | 80.0 [0.72, 0.87] | 0.37 [0.16, 0.83] | 0.02 |
|  | 75 or older | 86 | 87.2 [0.78, 0.93] | 0.61 [0.28, 1.37] | 0.24 |
| Ethnicity | white | 59 | 84.3 [0.81, 0.87] | Reference | . |
|  | Indigenous | 192 | 91.5 [0.81, 0.97] | 1.62 [1.23, 2.15] | <0.001 |
|  | Other | 744 | 88.5 [0.83, 0.93] | 1.11 [0.83, 1.48] | 0.5 |
|  | NA | 20 | 80.0 [0.56, 0.94] | 0.56 [0.12, 2.51] | 0.44 |
| Region | Urban | 347 | 88.5 [0.85, 0.92] | Reference | . |
|  | Suburban E | 195 | 84.6 [0.79, 0.89] | 0.92 [0.8, 1.06] | 0.27 |
|  | Suburban S | 197 | 85.8 [0.80, 0.90] | 0.84 [0.77, 0.91] | <0.001 |
|  | Suburban W | 139 | 82.7 [0.75, 0.89] | 0.71 [0.66, 0.77] | <0.001 |
|  | Rural | 137 | 81.0 [0.73, 0.87] | 0.76 [0.62, 0.93] | 0.01 |
| Income | < \$20,000 | 51 | 92.2 [0.81, 0.98] | Reference | . |
| | \$20,000 to \$39,999 | 72 | 88.9 [0.79, 0.95] | 1.00 [0.37, 2.74] | 1 |
| | \$40,000 to \$59,999 | 102 | 85.3 [0.77, 0.92] | 0.70 [0.25, 1.91] | 0.48 |
| | \$60,000 to \$79,999 | 125 | 85.6 [0.78, 0.91] | 0.63 [0.34, 1.16] | 0.14 |
| | \$80,000 to \$99,999 | 162 | 82.7 [0.76, 0.88] | 0.47 [0.24, 0.93] | 0.03 |
| | \$100,000 to \$119,000 | 141 | 86.5 [0.80, 0.92] | 0.64 [0.33, 1.26] | 0.2 |
| | \$120,000 or more | 276 | 82.6 [0.78, 0.87] | 0.55 [0.24, 1.27] | 0.16 |
|  | NA | 86 | 90.7 [0.82, 0.96] | 1.23 [0.39, 3.82] | 0.73 |
| **Forest** | 25+ per year | 145 | 89.7 [0.84, 0.94] | Reference | . |
|  | 11-25 per year | 226 | 87.6 [0.83, 0.92] | 0.77 [0.4, 1.48] | 0.44 |
|  | 2-10 per year | 302 | 87.1 [0.83, 0.91] | 0.72 [0.41, 1.26] | 0.25 |
|  | 2/year | 231 | 80.5 [0.75, 0.85] | 0.47 [0.31, 0.73] | <0.001 |
|  | never | 111 | 81.1 [0.73, 0.88] | 0.47 [0.21, 1.05] | 0.06 |
| **Environmentally Applied: Biological Acceptability** |  |  |  |  |  |
|  | **Category** | **No. respondents in category** | **Proportion of respondents with high acceptability [95%CI]** | **OR [95%CI]** | **p-value** |
| **Gender** | Woman | 532 | 74.6 [0.71, 0.78] | Reference | . |
|  | Male | 474 | 81.0 [0.77, 0.84] | 1.32 [0.92, 1.89] | 0.14 |
|  | Non-Binary | 8 | 37.5 [0.09, 0.76] | 0.17 [0.09, 0.32] | <0.001 |
|  | Other | 1 | 100.0 [0.03, 1.00] | . | . |
| **Age** | 18 to 24 | 128 | 86.7 [0.80, 0.92] | Reference | . |
|  | 25 to 34 | 185 | 74.6 [0.68, 0.81] | 0.37 [0.2, 0.7] | <0.001 |
|  | 35 to 44 | 180 | 75.6 [0.69, 0.82] | 0.35 [0.27, 0.46] | <0.001 |
|  | 45 to 54 | 153 | 76.5 [0.69, 0.83] | 0.41 [0.29, 0.59] | <0.001 |
|  | 55 to 64 | 168 | 75.0 [0.68, 0.81] | 0.38 [0.23, 0.61] | <0.001 |
|  | 65 to 74 | 115 | 76.5 [0.68, 0.84] | 0.41 [0.23, 0.72] | <0.001 |
|  | 75 or older | 86 | 80.2 [0.70, 0.88] | 0.47 [0.2, 1.07] | 0.07 |
| **Region** | Urban | 347 | 79.8 [0.75, 0.84] | Reference | . |
|  | Suburban E | 195 | 83.6 [0.78, 0.88] | 1.33 [1, 1.75] | 0.05 |
|  | Suburban S | 197 | 73.1 [0.66, 0.79] | 0.74 [0.64, 0.87] | <0.001 |
|  | Suburban W | 139 | 73.4 [0.65, 0.81] | 0.78 [0.59, 1.03] | 0.08 |
|  | Rural | 137 | 72.3 [0.64, 0.80] | 0.72 [0.54, 0.96] | 0.03 |
| **Income** | < \$20,000 | 51 | 68.6 [0.54, 0.81] | Reference | . |
| | \$20,000 to \$39,999 | 72 | 83.3 [0.73, 0.91] | 2.68 [1.41, 5.07] | <0.001 |
| | \$40,000 to \$59,999 | 102 | 77.5 [0.68, 0.85] | 1.80 [0.61, 5.36] | 0.29 |
| | \$60,000 to \$79,999 | 125 | 75.2 [0.67, 0.82] | 1.35 [0.72, 2.51] | 0.35 |
| | \$80,000 to \$99,999 | 162 | 83.3 [0.77, 0.89] | 1.76 [0.64, 4.82] | 0.27 |
| | \$100,000 to \$119,000 | 141 | 80.9 [0.73, 0.87] | 1.68 [0.58, 4.81] | 0.34 |
| | \$120,000 or more | 276 | 76.1 [0.71, 0.81] | 1.50 [0.92, 2.45] | 0.11 |
|  | NA | 86 | 67.4 [0.56, 0.77] | 1.13 [0.49, 2.64] | 0.77 |
| **Education** | Highschool or less | 130 | 73.1 [0.65, 0.80] | Reference | . |
|  | College | 288 | 78.8 [0.74, 0.83] | 1.42 [0.81, 2.49] | 0.22 |
|  | University + | 594 | 77.6 [0.74, 0.81] | 1.30 [1.05, 1.6] | 0.02 |
|  | Other/ NA | 3 | 66.7 [0.09, 0.99] | 1.11 [0.13, 9.4] | 0.93 |
| **Language** | English | 839 | 76.0 [0.73, 0.79] | . | . |
|  | French | 137 | 85.4 [0.78, 0.91] | 1.90 [1.04, 3.47] | 0.04 |
|  | Other | 39 | 76.9 [0.61, 0.89] | 0.96 [0.66, 1.41] | 0.85 |
| **Occupation** | Indoors | 591 | 76.1 [0.72, 0.80] | Reference | . |
|  | Mixed | 128 | 79.7 [0.72, 0.86] | 0.81 [0.44, 1.48] | 0.49 |
|  | Outdoors | 37 | 81.1 [0.65, 0.92] | 0.80 [0.54, 1.18] | 0.26 |
|  | Other | 259 | 78.4 [0.73, 0.83] | 1.25 [1.11, 1.4] | <0.001 |
| **Tick encounter** | Never | 782 | 73.8 [0.71, 0.77] | Reference | . |
|  | 1-2 times | 183 | 89.6 [0.84, 0.94] | 2.25 [1.12, 4.5] | 0.02 |
|  | 3+ times | 50 | 88.0 [0.76, 0.95] | 1.47 [0.33, 6.62] | 0.61 |
| **Landscape Management Acceptability** |  |  |  |  |  |
|  | **Category** | **No. respondents in category** | **Proportion of respondents with high acceptability [95%CI]** | **OR [95%CI]** | **p-value** |
| **Ethnicity** | white | 59 | 87.1 [0.84, 0.89] | Reference | . |
|  | Indigenous | 192 | 93.2 [0.84, 0.98] | 2.92 [1.33, 6.42] | 0.01 |
|  | Other | 744 | 82.8 [0.77, 0.88] | 0.81 [0.62, 1.05] | 0.11 |
|  | NA | 20 | 60.0 [0.36, 0.81] | 0.19 [0.05, 0.68] | 0.01 |
| **Region** | Urban | 347 | 85.6 [0.81, 0.89] | Reference | . |
|  | Suburban E | 195 | 88.2 [0.83, 0.92] | 1.09 [0.91, 1.3] | 0.37 |
|  | Suburban S | 197 | 82.7 [0.77, 0.88] | 0.65 [0.61, 0.71] | <0.001 |
|  | Suburban W | 139 | 89.9 [0.84, 0.94] | 1.31 [1.12, 1.53] | <0.001 |
|  | Rural | 137 | 85.4 [0.78, 0.91] | 0.84 [0.73, 0.96] | 0.01 |
| **Income** | < \$20,000 | 51 | 68.6 [0.54, 0.81] | Reference | . |
| | \$20,000 to \$39,999 | 72 | 80.6 [0.70, 0.89] | 1.85 [0.84, 4.05] | 0.13 |
| | \$40,000 to \$59,999 | 102 | 91.2 [0.84, 0.96] | 5.00 [1.94, 12.9] | <0.001 |
| | \$60,000 to \$79,999 | 125 | 88.0 [0.81, 0.93] | 3.06 [1.51, 6.19] | <0.001 |
| | \$80,000 to \$99,999 | 162 | 86.4 [0.80, 0.91] | 2.04 [0.67, 6.24] | 0.21 |
| | \$100,000 to \$119,000 | 141 | 88.7 [0.82, 0.93] | 2.58 [1.17, 5.7] | 0.02 |
| | \$120,000 or more | 276 | 85.1 [0.80, 0.89] | 1.75 [1.09, 2.81] | 0.02 |
|  | NA | 86 | 90.7 [0.82, 0.96] | 3.98 [1.52, 10.45] | 0.01 |
| **Yard** | No access | 171 | 83.6 [0.77, 0.89] | Reference | . |
|  | Access, no maintenance | 250 | 81.2 [0.76, 0.86] | 0.91 [0.67, 1.23] | 0.53 |
|  | Access and maintenance | 594 | 88.9 [0.86, 0.91] | 1.69 [1.18, 2.42] | <0.001 |
| **Pet** | Yes | 393 | 83.2 [0.79, 0.87] | Reference | . |
|  | No | 622 | 87.9 [0.85, 0.90] | 1.93 [1.32, 2.84] | <0.001 |
| **Host Targeted Methods: Small Mammal Acceptability** |  |  |  |  |  |
|  | **Category** | **No. respondents in category** | **Proportion of respondents with high acceptability [95%CI]** | **OR [95%CI]** | **p-value** |
| **Ethnicity** | white | 59 | 83.1 [0.71, 0.92] | Reference | . |
|  | Indigenous | 192 | 76.0 [0.69, 0.82] | 1.75 [1.27, 2.4] | <0.001 |
|  | Other | 744 | 74.9 [0.72, 0.78] | 1.12 [0.88, 1.41] | 0.36 |
|  | NA | 20 | 75.0 [0.51, 0.91] | 1.06 [0.74, 1.53] | 0.75 |
| **Region** | Urban | 347 | 77.5 [0.73, 0.82] | Reference | . |
|  | Suburban E | 195 | 76.9 [0.70, 0.83] | 0.94 [0.78, 1.12] | 0.48 |
|  | Suburban S | 197 | 73.1 [0.66, 0.79] | 0.82 [0.77, 0.87] | <0.001 |
|  | Suburban W | 139 | 72.7 [0.64, 0.80] | 0.75 [0.65, 0.87] | <0.001 |
|  | Rural | 137 | 75.2 [0.67, 0.82] | 0.85 [0.72, 1] | 0.05 |
| **Residence in Canada** | Less than 1 year | 13 | 92.3 [0.64, 1.00] | Reference | . |
|  | 1-5 years | 45 | 68.9 [0.53, 0.82] | 0.20 [0.05, 0.73] | 0.02 |
|  | 6-10 years | 37 | 83.8 [0.68, 0.94] | 0.56 [0.06, 5.23] | 0.61 |
|  | 10+ years | 920 | 75.3 [0.72, 0.78] | 0.38 [0.12, 1.16] | 0.09 |
| **Education** | Highschool or less | 130 | 68.5 [0.60, 0.76] | Reference | . |
|  | College | 288 | 76.4 [0.71, 0.81] | 1.63 [1.03, 2.57] | 0.04 |
|  | University + | 594 | 76.8 [0.73, 0.80] | 1.54 [1, 2.37] | 0.05 |
|  | Other/ NA | 3 | 66.7 [0.09, 0.99] | 0.55 [0.04, 7.88] | 0.66 |
| **Language** | English | 839 | 74.5 [0.71, 0.77] | Reference | . |
|  | French | 137 | 77.4 [0.69, 0.84] | 1.18 [0.85, 1.63] | 0.33 |
|  | Other | 39 | 92.3 [0.79, 0.98] | 3.85 [1.44, 10.33] | 0.01 |
| **Tick encounter** | Never | 782 | 72.8 [0.69, 0.76] | Reference | . |
|  | 1-2 times | 183 | 86.3 [0.80, 0.91] | 2.63 [1.36, 5.09] | <0.001 |
|  | 3+ times | 50 | 80.0 [0.66, 0.90] | 1.48 [0.4, 5.52] | 0.56 |
| **Host Targeted Methods: Deer Acceptability** |  |  |  |  |  |
|  | **Category** | **No. respondents in category** | **Proportion of respondents with high acceptability [95%CI]** | **OR [95%CI]** | **p-value** |
| **Gender** | Woman | 532 | 63.7 [0.59, 0.68] | Reference | . |
|  | Male | 474 | 65.4 [0.61, 0.70] | 0.98 [0.67, 1.43] | 0.93 |
|  | Non-Binary | 8 | 87.5 [0.47, 1.00] | 3.93 [2.01, 7.71] | <0.001 |
|  | Other | 1 | 0.0 [0.00, 0.98] | . | . |
| **Ethnicity** | white | 59 | 79.7 [0.67, 0.89] | Reference | . |
|  | Indigenous | 192 | 64.1 [0.57, 0.71] | 2.05 [0.91, 4.61] | 0.08 |
|  | Other | 744 | 64.0 [0.60, 0.67] | 0.96 [0.67, 1.39] | 0.84 |
|  | NA | 20 | 50.0 [0.27, 0.73] | 0.59 [0.43, 0.8] | <0.001 |
| **Region** | Urban | 347 | 67.7 [0.63, 0.73] | Reference | . |
|  | Suburban E | 195 | 69.7 [0.63, 0.76] | 1.14 [1.05, 1.24] | <0.001 |
|  | Suburban S | 197 | 64.5 [0.57, 0.71] | 0.90 [0.82, 0.99] | 0.03 |
|  | Suburban W | 139 | 61.9 [0.53, 0.70] | 0.85 [0.76, 0.94] | <0.001 |
|  | Rural | 137 | 52.6 [0.44, 0.61] | 0.55 [0.49, 0.62] | <0.001 |
| **Education** | Highschool or less | 130 | 54.6 [0.46, 0.63] | Reference | . |
|  | College | 288 | 66.7 [0.61, 0.72] | 1.71 [1.35, 2.17] | <0.001 |
|  | University + | 594 | 65.8 [0.62, 0.70] | 1.67 [1.14, 2.46] | 0.01 |
|  | Other/ NA | 3 | 66.7 [0.09, 0.99] | 1.62 [0.15, 17.93] | 0.69 |
| **Forest** | 25+ per year | 145 | 60.0 [0.52, 0.68] | Reference | . |
|  | 11-25 per year | 226 | 66.4 [0.60, 0.73] | 1.20 [1.04, 1.38] | 0.01 |
|  | 2-10 per year | 302 | 67.9 [0.62, 0.73] | 1.46 [0.98, 2.16] | 0.06 |
|  | 2/year | 231 | 62.8 [0.56, 0.69] | 1.21 [0.81, 1.81] | 0.35 |
|  | never | 111 | 62.2 [0.52, 0.71] | 1.30 [0.69, 2.44] | 0.42 |
| **Tick encounter** | Never | 782 | 62.0 [0.59, 0.65] | Reference | . |
|  | 1-2 times | 183 | 75.4 [0.69, 0.81] | 1.85 [1.46, 2.34] | <0.001 |
|  | 3+ times | 50 | 66.0 [0.51, 0.79] | 1.10 [0.3, 4] | 0.88 |
| **Personal Prevention: Behavioral Acceptability** |  |  |  |  |  |
|  | **Category** | **No. respondents in category** | **Proportion of respondents with high acceptability [95%CI]** | **OR [95%CI]** | **p-value** |
| **Gender** | Woman | 532 | 96.8 [0.95, 0.98] | Reference | . |
|  | Male | 474 | 92.4 [0.90, 0.95] | 0.35 [0.19, 0.67] | <0.001 |
|  | Non-Binary | 8 | 100.0 [0.63, 1.00] | . | . |
|  | Other | 1 | 100.0 [0.03, 1.00] | . | . |
| **Region** | Urban | 347 | 94.5 [0.92, 0.97] | Reference | . |
|  | Suburban E | 195 | 96.9 [0.93, 0.99] | 2.52 [2.13, 2.97] | <0.001 |
|  | Suburban S | 197 | 91.9 [0.87, 0.95] | 0.67 [0.51, 0.86] | <0.001 |
|  | Suburban W | 139 | 95.7 [0.91, 0.98] | 0.82 [0.42, 1.59] | 0.55 |
|  | Rural | 137 | 95.6 [0.91, 0.98] | 1.06 [0.65, 1.74] | 0.82 |
| **Language** | English | 839 | 95.5 [0.94, 0.97] | Reference | . |
|  | French | 137 | 89.8 [0.83, 0.94] | 0.25 [0.12, 0.51] | <0.001 |
|  | Other | 39 | 97.4 [0.87, 1.00] | 2.26 [0.42, 12.24] | 0.35 |
| **Tick bite** | Never | 848 | 95.2 [0.93, 0.97] | Reference | . |
|  | 1-2 times | 148 | 93.9 [0.89, 0.97] | 0.17 [0.05, 0.65] | 0.01 |
|  | 3+ times | 19 | 84.2 [0.60, 0.97] | 0.06 [0.02, 0.19] | <0.001 |
| **Tick encounter** | Never | 782 | 94.1 [0.92, 0.96] | Reference | . |
|  | 1-2 times | 183 | 96.7 [0.93, 0.99] | 11.17 [1.5, 83.29] | 0.02 |
|  | 3+ times | 50 | 98.0 [0.89, 1.00] | 37.84 [1.12, 1275.63] | 0.04 |
| **Personal Prevention: Chemical Acceptability** |  |  |  |  |  |
|  | **Category** | **No. respondents in category** | **Proportion of respondents with high acceptability [95%CI]** | **OR [95%CI]** | **p-value** |
| **Age** | 18 to 24 | 128 | 96.1 [0.91, 0.99] | Reference | . |
|  | 25 to 34 | 185 | 87.6 [0.82, 0.92] | 0.34 [0.09, 1.33] | 0.12 |
|  | 35 to 44 | 180 | 87.2 [0.81, 0.92] | 0.29 [0.07, 1.2] | 0.09 |
|  | 45 to 54 | 153 | 85.6 [0.79, 0.91] | 0.26 [0.07, 0.97] | 0.05 |
|  | 55 to 64 | 168 | 84.5 [0.78, 0.90] | 0.20 [0.06, 0.68] | 0.01 |
|  | 65 to 74 | 115 | 82.6 [0.74, 0.89] | 0.16 [0.05, 0.51] | <0.001 |
|  | 75 or older | 86 | 94.2 [0.87, 0.98] | 0.51 [0.08, 3.43] | 0.49 |
| **Region** | Urban | 347 | 86.5 [0.82, 0.90] | Reference | . |
|  | Suburban E | 195 | 90.3 [0.85, 0.94] | 1.83 [1.64, 2.04] | <0.001 |
|  | Suburban S | 197 | 89.3 [0.84, 0.93] | 1.39 [1.12, 1.73] | <0.001 |
|  | Suburban W | 139 | 85.6 [0.79, 0.91] | 0.81 [0.64, 1.02] | 0.07 |
|  | Rural | 137 | 87.6 [0.81, 0.93] | 1.21 [0.96, 1.53] | 0.12 |
| **Language** | English | 839 | 88.6 [0.86, 0.91] | Reference | . |
|  | French | 137 | 81.8 [0.74, 0.88] | 0.46 [0.28, 0.76] | <0.001 |
|  | Other | 39 | 92.3 [0.79, 0.98] | 2.30 [0.83, 6.4] | 0.11 |
| **Tick encounter** | Never | 782 | 86.1 [0.83, 0.88] | Reference | . |
|  | 1-2 times | 183 | 92.3 [0.87, 0.96] | 1.74 [1.23, 2.48] | <0.001 |
|  | 3+ times | 50 | 98.0 [0.89, 1.00] | 6.40 [1.49, 27.53] | 0.01 |
| **Lyme disease diagnosis** | Yes | 34 | 91.2 [0.76, 0.98] | Reference | . |
|  | No | 967 | 87.9 [0.86, 0.90] | 1.17 [0.73, 1.89] | 0.51 |
|  | Unknown | 14 | 71.4 [0.42, 0.92] | 0.16 [0.06, 0.44] | <0.001 |

For landscape management strategies, Indigenous respondents had higher odds of finding these to be acceptable when compared to white respondents (OR: 2.92; 95% CI: 1.33, 6.42; p-value <0.05). Individuals residing in the Suburban West area had 1.31 higher odds of acceptability for landscape management strategies when compared to those in the Urban center (95% CI: 1.12, 1.53; p-value<0.001). Respondents who had access to a yard and were responsible for maintaining it had higher odds of acceptability for these strategies when compared to those with no access to a yard (OR: 1.69; 95% CI: 1.18, 2.42; p-value<0.001) (Table 7).

Within the host-targeted strategy category, Indigenous respondents had higher odds of finding small mammal-targeted strategies to be acceptable (OR: 1.75; 95% CI: 1.27, 2.40; p-value <0.001) when compared to white respondents. Most individuals living outside of the Urban center (excluding Suburban East) had significantly lower acceptability with ORs ranging from 0.75 to 0.85 (p-values all <0.05). Similarly for deer targeted strategies, region played an important role in acceptability with those residing in Suburban South, Suburban West, and Rural areas having lower acceptability of deer targeted strategies (OR: 0.91; 95% CI: 0.82, 0.99; p-value <0.05, OR: 0.85; 95% CI: 0.76, 0.94; p-value<0.001, OR: 0.55, 95% CI: 0.49, 0.62; p-value <0.001) respectively, and those living in Suburban East having slightly higher odds of acceptability (OR: 1.14; 95% CI: 1.05, 1.24; p-value<0.001) (Table 7).

We observed similar patterns across behavioural and chemical based personal prevention strategies. Residents of Suburban East had higher odds of high acceptability of these strategies compared with urban residents (behavioural: OR: 2.52; 95% CI: 2.13–2.97; p<0.001; chemical: OR: 1.83; 95% CI: 1.64–2.04; p<0.001). Additionally, respondents who identified as primarily French-speaking had lower odds of high acceptability than English-speaking respondents for behavioural and chemical personal protection strategies respectively (OR: 0.25; 95% CI: 0.12– 0.51; p<0.001; OR: 0.46; 95% CI: 0.28–0.76; p<0.001). Finally, prior tick encounters were also associated with greater odds of high acceptability for both personal prevention strategy types when compared to individuals with no encounters (behavioural: ORs of 11.17 to 37.84; p-values <0.02 to 0.04) (chemical: ORs: 1.74 to 6.40; p-values <0.001 to <0.05) (Table 7).

#### 3.2.3 Synthesis across intervention specific models

Figure 3 summarizes the patterns in variables that were most consistently included and significantly associated with intervention acceptability. Across intervention-specific models, region, gender, and age were the significant sociodemographic predictors of acceptability that were most consistently retained following DAG and AIC based model refinement, with length of residence in Canada, and language being less likely to be retained or significant in the models (Fig. 3). Risk and exposure variables showed greater variability across interventions, with occupation and tick encounter frequency being the most frequently retained and associated with acceptability, whereas both suspected and confirmed Lyme disease diagnosis were never, or rarely included in the models.

**Figure 3:**
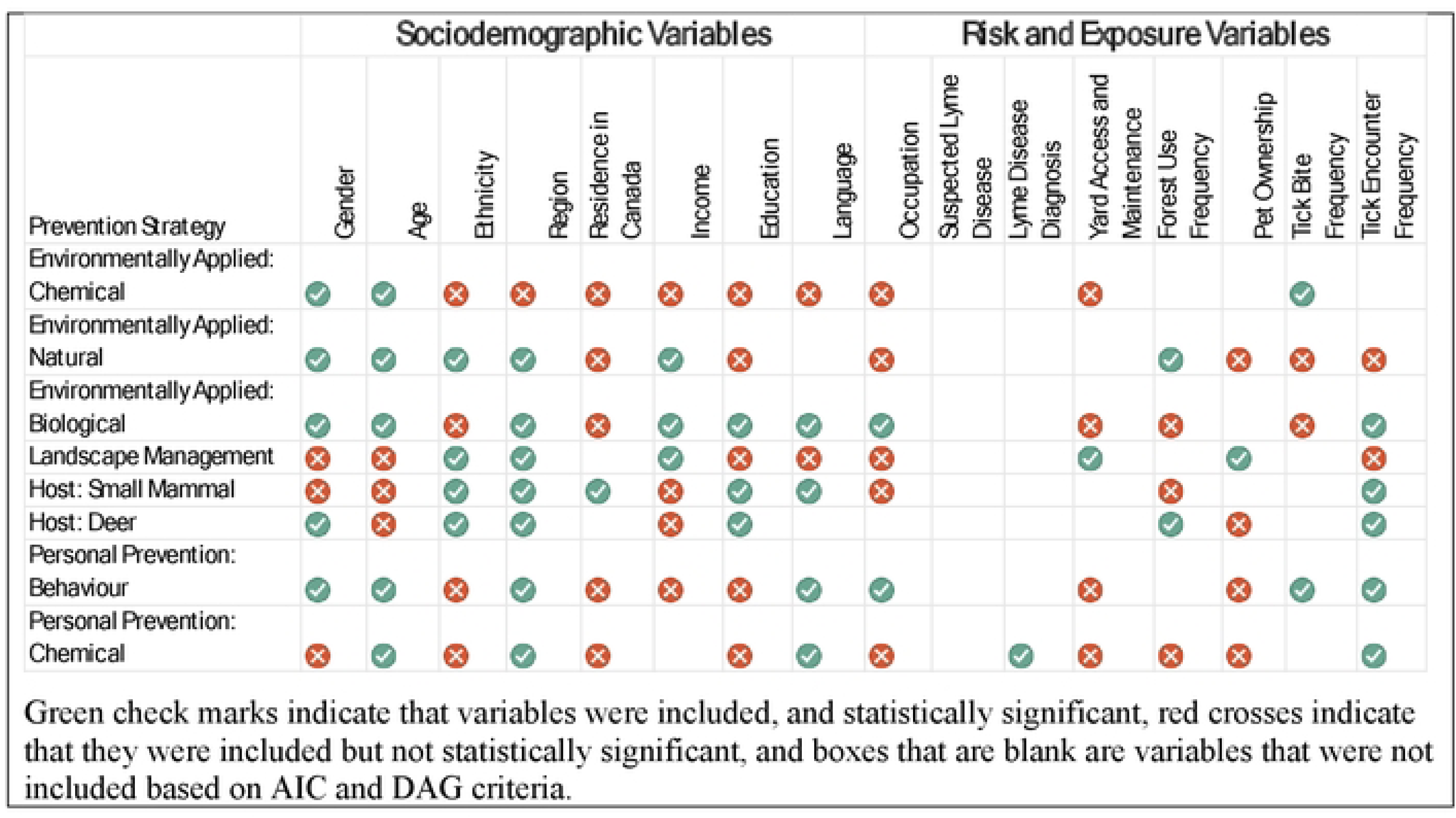
Overview of Variables Included in Each Multivariable Model and Their Statistical Significance Across Eight Acceptability Domains

#### 3.2.4 Domain-specific model comparison using AIC of factors associated with high acceptability of intervention categories

Sixteen domain-specific models (two per intervention: sociodemographic-only and risk/exposure-only) were estimated to compare relative contributions to model fit. Across interventions, patterns of AIC change differed between sociodemographic and risk and exposure models, suggesting that the relative contribution of variables to model fit varies by intervention type (Supplemental Table 3). This approach identified sociodemographic factors as being more consistently associated with acceptability of interventions compared to risk and exposure factors. In particular, the sociodemographic factors related to gender, region of residence, age, and income were the most frequently associated with moderate to high model fit contributions.

Among risk and exposure variables, occupation, yard management, forest visitation, pet ownership, and tick encounters, contributed to better model fit most consistently.

#### 3.2.5 Core Generalized estimating equation (GEE) model results

Variables included in the forced core model were variables that changed the AIC by more than two points in more than three of the models or were informed by the DAG. This process resulted in the final inclusion of sociodemographic variables (gender, age group, ethnicity, region of residence, length of residence in Canada, income and education level) and risk or exposure variables (outdoor occupation, tick encounter frequency in the last year, and forest visitation frequency) in the Generalized estimating equation (GEE) model. Both personal prevention approaches (behavioural approach and chemical approach) had the highest probability of having high acceptability (0.95, 95% CI: 0.93, 0.96, p-value<0.001, and 0.88, 95% CI: 0.86, 0.90, p-value<0.001), respectively (Figure 4). Deer targeted interventions and environmentally applied chemical strategies had the lowest probability of acceptability (0.65, 95% CI: 0.62, 0.68, p-value <0.001) and 0.57%, 95% CI: 0.54, 0.60, p-value <0.001), respectively (Fig. 4).

**Figure 4:**
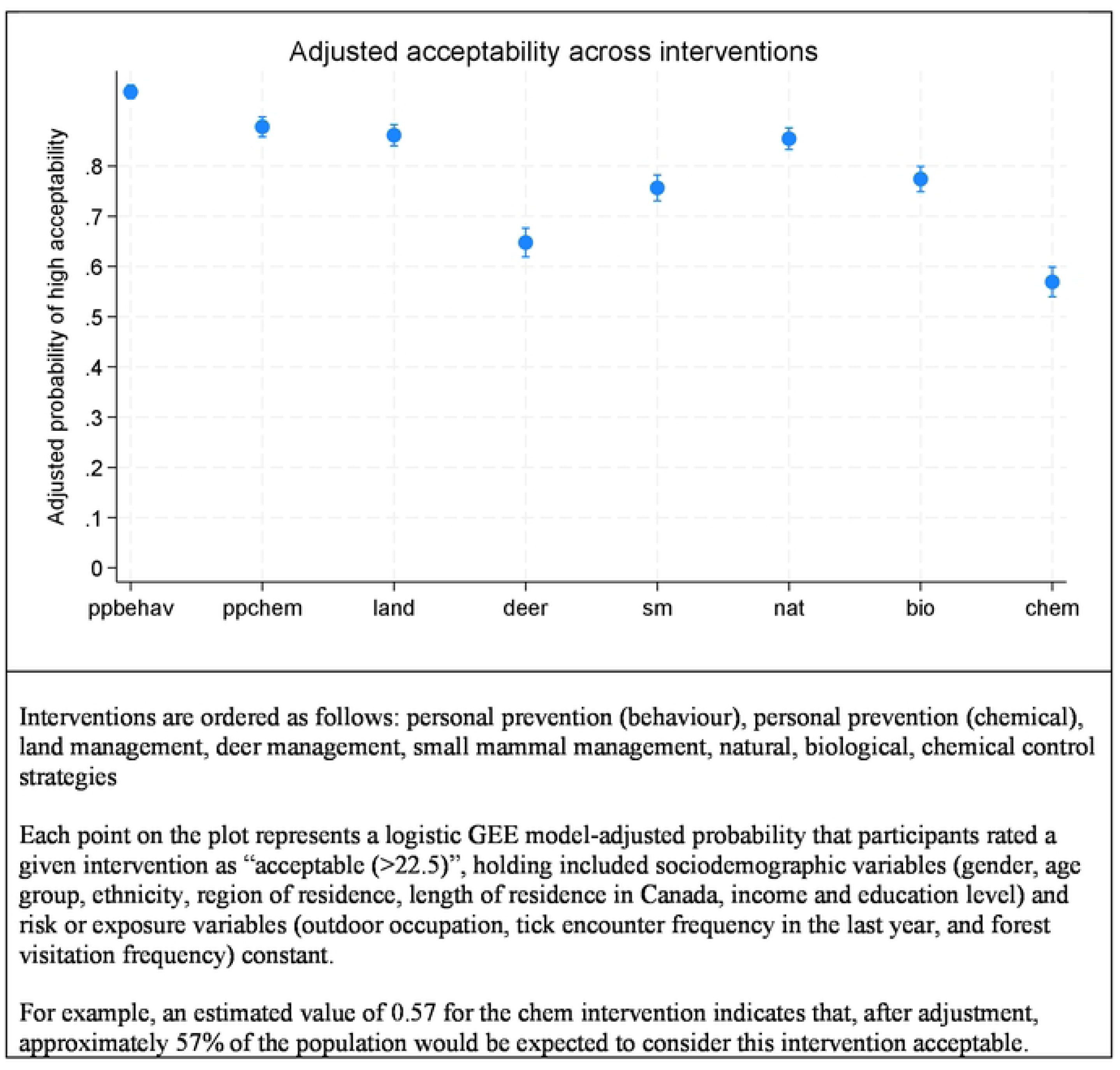
Model-adjusted probability of high acceptability across Lyme disease prevention and tick control interventions

## 4. Discussion

In the context of increasing public health risks posed by LD in Ottawa, identifying prevention and control strategies that are both effective and socially acceptable are central to increasing the uptake and sustainability of interventions (34). This exploratory sequential mixed methods study demonstrated that overall levels of acceptability of Lyme disease prevention and tick control interventions among Ottawa adults was high, with the proportion reporting high acceptability ranging from 57% to 95% across eight intervention types. Acceptability varied across these intervention types. Personal protection strategies (both chemical and behavioural) were consistently rated as highly acceptable according to both the key informant interviews (KIIs) and survey results, whereas chemical control approaches and deer targeted host management strategies were among the least acceptable. Landscape management strategies were generally acceptable, with the exception of vegetation burning, as identified in the KIIs. Importantly, patterns of acceptability were not uniform across the population.

Region of residence was the most consistently retained sociodemographic predictor. Residents of the Suburban East were more likely to have high acceptability scores for most interventions when compared to urban residents. This pattern was observed across all intervention types, except for small mammal-targeted strategies and natural alternatives to synthetic acaricides.

Furthermore, we observed differences in acceptability across gender, age, ethnicity, and language groups. While direct comparisons are limited by differences in measurement approaches, our findings contrast with acceptability patterns in other Canadian contexts, underscoring the importance of local context in shaping public support for LD prevention strategies (20). Tick encounter frequency was the risk and exposure variable most frequently retained in the individual models. Individuals with one to two self-reported tick encounters in the last year had higher odds of having high acceptability of interventions compared to individuals with no self-reported tick encounter. This association was the strongest for chemical and behavioural personal prevention strategies. Tick bite frequency, forest use frequency, and occupation, were also frequently included in individual models, however they were less likely to be associated with acceptability than sociodemographic variables. This may suggest that acceptability may depend more on social and geographic context than by individual risk exposure alone.

Importantly, these findings also highlight important differences between knowledge, attitudes, and practices and acceptability, even within the same regional context. In a previous KAP study conducted in Ottawa, older age, yard characteristics, exposure index, and knowing someone with LD were key drivers of knowledge, while prevention practices were most strongly associated with non-Indigenous racialized identity, yard access, previous LD or tick exposure, and occupation (indoor versus outdoor) (28). In contrast, our findings suggest that the acceptability of tick and LD interventions is more consistently associated with sociodemographic factors compared to risk or exposure characteristics. These differences are important to consider in the design of implementation strategies and public outreach surrounding interventions. While previous studies have identified risk and exposure variables such as occupation, recreational activities, outdoor space use, pet ownership, and home or yard characteristics, and tick encounters or bites have been widely documented as contributors to LD risk, their influence on acceptability in our study was more intervention-specific (35). This suggests that perceived risk alone may not consistently translate into support for all prevention strategies, and that acceptability is shaped by how individuals perceive feasibility and consequences of specific interventions in context. These findings were also supported by our qualitative findings, which indicated that individuals considered a wide range of factors while thinking about acceptability of a given intervention, and were particularly concerned with feasibility, and unintended consequences of choosing a certain intervention type (36,37).

The qualitative phase of this study was instrumental in shaping both the survey instrument and the interpretation of quantitative findings. Key informant interviews highlighted themes such as those mentioned above, along with responsibility for implementation, and the scale of the intervention, which informed the refinement of survey questions to ensure clarity and consistent interpretation by the general public. These interviews also underscored the importance of providing brief educational explanations for each intervention, we therefore opted to include brief infographic-style descriptions in the survey to help establish a base understanding of these interventions in a quick and digestible way, with the aim of increasing comprehension and reducing misinterpretation of potentially unfamiliar intervention options. Additionally, qualitative findings consistently supported our quantitative findings, as many of the themes from the key informants were also represented in the quantitative results, and the relative acceptability of the interventions was also echoed between the interview and survey results. This convergence of results was a strength of this study, as we were able to improve our confidence in the validity of the acceptability patterns observed while using an acceptability scale novel to this context and use.

In terms of preventive behaviours in our study population in Ottawa, Ontario, although respondents reported a high level of tick encounters and bites (22% and 16% respectively), only 10% consistently used personal prevention measures, and 21.7% reported using yard or environmental controls. In contrast, previous research in another high risk locale in the United States noted that 99% of survey respondents reported using personal protective behaviours, and 65% reported using environmental tick controls (21). Our findings for personal protection are more consistent with other research performed in the neighbouring province of Quebec, where authors found that while tick exposure in the past year ranged from 3.4 to 21.9% (defined as reported encounter of at least one tick in the last year), personal preventive behaviours varied from 27.0% to 30.1% (38). Differences in environmental conditions, exposure patterns, and population familiarity with interventions all contribute to variations in acceptability across settings, highlighting the need for context specific evaluations when informing decision making for this health issue.

There were several limitations that should be considered when interpreting these findings. We examined distributions of summed acceptability scores to assess their overall shape and range. Although distributions of summed acceptability scores were approximately normal, analysis focused on a binary high versus low acceptability outcome to support interpretation in an implementation context. While dichotomization of the outcome variables may oversimplify the complexity of these relationships, it facilitates interpretation by stakeholders and enabled consistent comparison across intervention types. Additionally, we performed a sensitivity analysis using alternative thresholds, which resulted in similar patterns, supporting the robustness of the primary results, and threshold score. Additionally, the leave one variable out approach based on changes in AIC reflects each predictor’s contribution to model fit conditional on the other variables included in the model (and should not be interpreted as a measure of independent or causal importance).

This study uses a novel approach to acceptability scoring within the context of LD prevention and tick control, which limits more direct comparability with some prior studies. However, the mixed methods nature of the study design provided validation of the score’s relevance for this field. The measures we included in the survey primarily focused on interventions over which individuals had some element of control over, rather than an external party.

Finally, the cross-sectional nature of this study prevents us from establishing a causal relationship between variables associated with high acceptability in the survey results. Data were collected through an online panel, and although the sample sociodemographic characteristics were descriptively compared to the most recent Canadian census data for Ottawa across age, sex, education, and income distributions, participation through a membership-based survey platform may introduce some element of selection bias, whereby the associations observed in the study may not fully reflect those in the target population.

Overall, these findings provide important insight into public support for LD prevention and tick control strategies across a range of urban classifications within a Canadian context and highlight the value of combining qualitative and quantitative approaches to inform feasible and contextually appropriate intervention planning, with high acceptability.

Recent cross-Canada qualitative research found that LD and tick bite prevention and adaptation efforts can influence mental health, outdoor recreation, interaction with nature, and views on pet ownership (39). These findings emphasize the importance of increasing education surrounding acceptable intervention options for control of this issue, while being cognisant of the stress and burden that tick-related risk can place on individuals and communities. Balancing risk with maintaining participation in outdoor spaces is challenging, but as shown by this, and previous studies performed in the Ottawa area, is critical to protecting human physical and mental health (39). Findings from this study indicate that personal prevention behaviours are highly acceptable and represent an accessible entry point for increased education and promotion of protective behaviours. However, there was also a high level of interest in landscape management options and environmentally applied natural alternatives to spraying chemicals to reduce tick abundance in human use areas. Together, these results provide a range of intervention options that can be implemented at both the individual and organizational or stakeholder level in the region.

## 5. Conclusion

In this study we linked key informant interview data with population level survey data. This study identified context specific factors that shape public acceptability of tick control and LD prevention interventions, providing data to better inform socially acceptable, feasible, and context relevant prevention strategies in the city of Ottawa.

## Data Availability

The data underlying this study are not publicly available due to ethical restrictions established by the informed consent agreements, which limit data access to the research team. These restrictions apply to both quantitative survey data and qualitative interview data. Requests for access to the data may be directed to the corresponding author, subject to institutional ethics review and approval.

## 6. Acknowledgements

Thank you to the participants who generously shared their time and experiences during the qualitative phase of this study. We would also like to thank Claudia Duguay and other members of the Threads Lab at Bruyere Research Institute for their time reviewing and testing survey materials during the survey pilot phase.

## CRediT Authors statement

Conceptualization: KO, KZ, AK, MK; Data curation: KO, MN; Formal analysis: KO, MN; Funding acquisition: KO, MK; Investigation: KO; Methodology: KO, KZ, CF, AK, MK; Project administration: KO; Supervision: AK, MK; Validation: CF, AK; Visualization: KO; Writing – original draft: KO; Writing – review & editing: KO, MN, KZ, CF, AK, MK.

